# Poor dream recall associates with Alzheimer’s disease biomarkers and dementia risk

**DOI:** 10.1101/2025.09.16.25335946

**Authors:** Darren M. Lipnicki, Meritxell Valentí, Elizabeth Valeriano-Lorenzo, Ashleigh S. Vella, María Ascensión Zea-Sevilla, Mario Ricciardi, Belén Frades, Minerva Martinez, Sonia Wagner, Perminder S. Sachdev, Teodoro del Ser, Pascual Sanchez-Juan

## Abstract

**INTRODUCTION:** Dreaming is subserved by the default mode network (DMN) and can be abolished by focal lesions within key regions. DMN dysfunction is promoted by pre-clinical Alzheimer’s pathology and apolipoprotein E (*APOE*) ε4 carriage, but any effects of these on dreaming are unknown.

**METHODS:** Associations between dream recall and *APOE* ε4 carriage and blood phosphorylated tau (p-tau) 217 levels were determined using data for 1049 cognitively normal adults. Associations between dream recall and cognitive decline and dementia risk were also investigated.

**RESULTS:** Higher p-tau217 levels and *APOE* ε4 carriage were both associated with lower chances of dream recall, independently of memory test scores and other covariates. Not recalling dreams at baseline was associated with faster cognitive decline and greater likelihood of dementia over 10 years of follow up.

**DISCUSSION:** Poor dream recall in later life may be a novel and easily assessed indicator of early neurodegeneration.

## 1. BACKGROUND

Dreams have fascinated people from all cultures since ancient times to the present day,^1^ but their recall varies among individuals^2^ and can be abolished by brain injury.^3^ Furthermore, a 1992 study found that lower dream recall frequency predicted incident dementia, an outcome considered “the least expected result” (p. 177) and not well understood at the time.^4^ However, current understanding of the default mode network (DMN), discovered in 2001,^5^ makes this result potentially explainable and relevant for identifying early signs of neurodegeneration.

The DMN comprises regions that subserve internally focused thought processes like self-reflection and mind wandering.^6,7^ The neurocognitive theory of dreaming posits that dreaming is also a form of internally generated thought subserved by the DMN.^8^ This is evidenced by neuroimaging and developmental studies, as well as lesions to DMN regions, including the medial prefrontal cortex (MPFC) and temporoparietal junction (TPJ), that abolish dreaming (reviewed in Domhoff^8^). Complementary studies of healthy young adults have reported that high dream recallers had greater regional cerebral blood flow than low dream recallers in the TPJ during rapid eye movement (REM) sleep, and in the TPJ and MPFC during both REM sleep and wakefulness.^9^ High dream recallers also had greater MPFC white matter density^10^ and increased functional connectivity between the MPFC and TPJ during waking rest.^11^

Individuals with Alzheimer’s disease (AD) show altered connectivity between DMN regions.^7^ These alterations develop pre-clinically and may identify individuals who will later exhibit cognitive decline^12^ or develop dementia.^13^ Amyloid beta (Aβ) reportedly first accumulates in the DMN,^14^ and has been associated with DMN hypoconnectivity in cognitively normal older adults,^14,15^ particularly under high tau-PET levels.^16,17^ Further, studies of patients with early atypical AD phenotypes have found that higher DMN tau levels were associated with lower connectivity^18^ and predicted clinical decline.^19^ Pre-clinical AD-like changes in DMN function have been associated with the apolipoprotein E (*APOE)* ε4 allele,^7,20,21^ the strongest genetic risk factor for sporadic AD.^22,23^ *APOE* ε4 carriage is associated with higher global Aβ and DMN tau among cognitively unimpaired older adults.^24,25^ Another study found disrupted DMN functionality in cognitively normal *APOE* ε4 carriers negative for Aβ, suggesting *APOE* ε4 may influence the DMN via additional pathways.^26^

Evidence suggests the DMN has a crucial role in dreaming, and reduced functionality is associated with less dream recall. Given DMN functionality is impaired in *APOE* ε4 carriers and by AD-related neuropathology pre-clinically, we hypothesized that dream recall will be reduced in cognitively normal older individuals who are *APOE* ε4 carriers or have relatively high plasma phosphorylated tau (p-tau) 217 levels, an accurate indicator of early AD pathology.^27^ We also expected less dream recall to be associated with future cognitive decline, and with dementia.^4^ Our analyses controlled for many factors recommended in dream recall studies, including memory test performance.^28^ Finding poor dream recall to be associated with *APOE* ε4 carriage, higher plasma p-tau217 levels, and future cognitive decline and dementia would support poor dream recall being a potential early manifestation of AD-related neurodegeneration.

## 2. METHODS

### 2.1 Participants

The Vallecas Project was a longitudinal cohort study (2011–2023) to identify early biomarkers of cognitive impairment in older adults.^29^ The participants were home-dwelling volunteers without relevant psychiatric or neurologic disorders or systemic disease, recruited through radio, TV and leaflet campaigns, and visits to centers for the elderly in Madrid, Spain. They undertook annual systematic assessments that included medical history, lifestyle habits, neurological and neuropsychological exams, and blood collection. From an initial sample of 1213, we excluded 164 individuals with mild cognitive impairment (MCI) or missing baseline data for dream recall, *APOE* ε4 status, p-tau217 or clinical diagnosis.

### 2.2 Dream recall

Dream recall was assessed by asking participants “Do you remember your dreams?” (yes/no).

### 2.3 *APOE* genotype and p-tau217

Plasma from baseline blood samples was isolated and stored at -80º. *APOE* genotype (rs429358 and rs7412) was determined by Real-Time PCR,^30^ and participants with ≥ 1 ε4 alleles were considered *APOE* ε4 carriers. Plasma p-tau217 levels were determined by the Fujirebio method,^31^ and > 0.247 pg/mL was considered high.^32^

### 2.4 Cognitive assessments

An extensive neuropsychological test battery was administered at every annual assessment. Five scores from three of the tests were used to obtain a modified Preclinical Alzheimer Cognitive Composite (PACCm)^33^ score for every participant at every assessment: free and total immediate recall and delayed free recall from the Free and Cued Selective Reminding Test (FCSRT),^34^ performance in 1 minute on the Digit-Symbol coding test,^35^ and Mini Mental State Examination (MMSE)^36^ score. Raw scores for these tests were transformed to z scores (using the baseline means and standard deviations [SDs] of cognitively normal participants) and averaged to form the PACCm score. For 11 participants without MMSE data at one assessment, a score was imputed based on scores for the prior and subsequent assessments.

### 2.5 Clinical diagnoses

A consensus diagnosis of cognitively normal, MCI or dementia was made by an experienced team (one neurologist and one neuropsychologist) for each participant at each assessment. Participants were considered cognitively normal when performance was above -1.5 SD of age- and education-based norms on all tests.

MCI was diagnosed according to the National Institute on Aging-Alzheimer’s Association criteria^37^ when: (1) the participant or informant reported problems to recall three or more of the following: talks, readings, names, places, plans, or orientation clues, impairment in daily activities due to memory failure, or concern about cognitive changes; (2) cognitive test performance was below -1.5 SD of norms for at least two of the following tests: immediate and delayed free recall of the FCSRT, immediate free recall of the Rey figure test, semantic and phonological verbal fluency, and time to copy for the Rey figure test; (3) instrumental activities of daily living were essentially preserved, as indicated by a Functional Assessment Questionnaire^38^ score < 6.

Dementia was diagnosed according to the Diagnostic and Statistical Manual of Mental Disorders, Fourth Edition, Text Revision (DSM-IV-TR)^39^ when (1) cognitive test performance was below -1.5 SD of norms for at least two tests, and (2) there were relevant functional deficits, as indicated by a Functional Assessment Questionnaire score ≥ 6.

The date of conversion to MCI or dementia was recorded as: a) the date of visit when the diagnosis was made in the Vallecas Project, or b) the date when the diagnosis was reported in the clinical records by the general practitioner. There were 28 cases of conversion to dementia among participants who did not regularly attend the planned visits, and for these the date of conversion to MCI was estimated as the midpoint between the last diagnosis of normal cognition and the first record of dementia.

### 2.6 Covariates

Demographic covariates were age, sex and education (less than or at least complete high school). Self-reported sleep-related covariates included insomnia (initial and maintenance), sleep duration (< 6 or ≥ 6 hours), awakenings per night, REM sleep behaviour disorder (strange movements, talking or making strange noises while sleeping), and restless legs syndrome (both tingling or pain in legs when sitting quietly or in bed and improvement after getting up and walking). Further self-reported covariates were current smoking, alcohol consumption, and use of medications including anxiolytics/hypnotics, anticholinergics, antidepressants, and dopaminergics. Additional covariates were Geriatric Depression Scale^40^ score, State-Trait Anxiety Inventory^41^ state and trait anxiety scores, and a Parkinsonism score (0–10 summing Parkinsonian gait, amimia, and resting tremor, bradykinesia, rigidity and cogwheel rigidity in right and left side).

### 2.7 Statistical analysis

Generalised linear models with a binomial distribution and a logit link function examined associations between dream recall status and each of *APOE* ε4 carriage, p-tau217 levels, and baseline cognitive performance. Adjusted models included baseline age (≤ 74 years = 0, > 74 years = 1, based on a median split) and sex (male = 0, female = 1), as well as *APOE* ε4 status when p-tau217 or cognitive performance were predictors and free delayed recall score when *APOE* ε4 status or p-tau217 were predictors. Also included were covariates significantly associated with dream recall in multiple logistic regressions controlling for age, sex, free delayed recall score and *APOE* ε4 status: initial insomnia, maintenance insomnia, sleep duration (≥ 6 hours), awakenings per night, and current smoking.

A linear mixed-effects model examined associations between dream recall status and longitudinal cognitive decline, controlling for sex, education, and *APOE* ε4 carriage. The model included a random intercept for each participant and a random slope for time to model individual differences in ageing effects. Fixed effects included time (age), dream recall status, time*dream recall status interaction, sex, and education. The model was fitted using restricted maximum likelihood, and Satterthwaite’s approximation was used for computing t-tests. Model assumptions were evaluated by inspecting residual distributions and random effect variances.

Cox proportional hazards regression models were employed to examine the association between dream recall status and conversion from normal cognition to MCI and to dementia. Univariate models first assessed the independent effects of dream recall status, age, sex, and *APOE* ε4 status. A subsequent multivariate model included dream recall, age and sex as simultaneous predictors. Since *APOE* ε4 carriage was highly related to dream recall, separate multivariate models with dream recall status, age, sex, were performed for *APOE* ε4 carriers and non-carriers. Proportional hazards assumptions were met for all included covariates.

Analyses were performed using R (version 4.2.3) with the *glm* function from the *stats* package, and the *lmer* function from the *lme4* and *lmerTest* packages. The *coxph* function, and *survival* and *survminer* packages were also used. *P* values were 2-sided, with α = 0.05.

## 3. RESULTS

### 3.1 Participant characteristics

Participant characteristics at baseline are summarised in Table 1 (the median [IQR] of continuous variables are additionally shown in Table S1). The sample’s mean age was 74.7 years and 64% were women. A total of 325 (31%) participants reported that they did not remember dreams. Compared to non-recallers, dream recallers were more likely to sleep ≥6 hours per night, have onset insomnia and more awakenings per night; they also had higher depression and trait anxiety scores. Conversely, non-recallers were more likely to have maintenance insomnia, and to be an *APOE* e4 carrier or current smoker; they were also older and had higher p-tau217 levels. Other factors including memory and PACCm scores and medication use were not related to dream recall (Table 1). The mean follow-up was 6.24 (SD = 3.98) years (range 0–11.5 years); 435 participants had a complete follow-up of 10 or more years. During follow-up, 208 participants (19.6%) developed incident MCI and 77 (7.3%) progressed to dementia.

**Table 1.** Participant characteristics at baseline.

| Characteristic | Total | Dream recall status |  | p value |
| --- | --- | --- | --- | --- |
|  | (n = 1049) | Yes (n = 724) | No (n = 325) |  |
| Demographics |  |  |  |  |
| Age, years | 74.7 (3.8) | 74.5 (3.8) | 75.1 (3.9) | 0.029 |
| Sex, female | 676 (64) | 480 (66) | 196 (60) | 0.061 |
| Education, <high school | 514 (49) | 368 (51) | 146 (45) | 0.077 |
| Biomarkers |  |  |  |  |
| APOE ε4 carrier | 172 (16) | 104 (14) | 68 (21) | 0.008 |
| p-tau217, pg/mL | 0.188 (0.547) | 0.185 (0.616) | 0.195 (0.334) | <0.001 |
| Cognitive test scores |  |  |  |  |
| Total delayed recall | 14.5 (1.7) | 14.5 (1.7) | 14.4 (1.7) | 0.791 |
| Free delayed recall | 9.6 (2.5) | 9.7 (2.5) | 9.3 (2.6) | 0.090 |
| PACCm, z-scores | 0 (0.7) | 0.02 (0.7) | -0.04 (0.8) | 0.208 |
| Covariates |  |  |  |  |
| Initial insomnia | 286 (27) | 214 (30) | 72 (22) | 0.013 |
| Maintenance insomnia | 298 (29) | 191 (27) | 107 (33) | 0.034 |
| Sleep duration, ≥6 hours | 855 (81) | 604 (83) | 251 (77) | 0.017 |
| Restless legs syndrome | 106 (10) | 69 (10) | 37 (12) | 0.391 |
| REM sleep behaviour disorder | 241 (24) | 170 (25) | 71 (24) | 0.764 |
| Awakenings per night | 1.5 (1.3) | 1.6 (1.3) | 1.36 | 0.036 |
| Parkinsonian symptoms | 0.2 (0.8) | 0.2 (0.7) | 0.2 (0.8) | 0.410 |
| Current smoking | 391 (37) | 249 (34) | 142 (44) | 0.004 |
| Alcohol drinker | 481 (46) | 326 (45) | 155 (48) | 0.423 |
| Anxiolytic/hypnotics | 422 (40) | 283 (39) | 139 (43) | 0.261 |
| Anticholinergics | 213 (20) | 139 (19) | 74 (23) | 0.184 |
| Antidepressants | 316 (30) | 222 (31) | 94 (29) | 0.570 |
| Dopaminergics | 163 (15) | 106 (15) | 57 (18) | 0.231 |
| GDS score | 1.6 (2.2) | 1.7 (2.3) | 1.3 (2.1) | 0.026 |
| State anxiety score | 14.4 (9) | 14.4 (8.9) | 14.4 (9.3) | 0.899 |
| Trait anxiety score | 16.9 (9.8) | 17.3 (9.8) | 16.1 (9.8) | 0.049 |

*Note:* Values are no. (%) for categorical variables and mean (SD) for continuous variables.
The median (IQR) of continuous variables is shown in Table S1.
Abbreviations: APOE, apolipoprotein E; GDS, Geriatric Depression Scale; PACCM, modified Preclinical Alzheimer Cognitive Composite; REM: rapid eye movement.

### 3.2 Associations between *APOE*ε4, p-tau217, and dream recall

*APOE* ε4 carriage and higher p-tau217 levels were both associated with a significantly lower likelihood of recalling dreams in the unadjusted models, and in the adjusted models that included age, sex, delayed free recall score and other covariates (Table 2). There was no association between PACCm scores and dream recall. A sensitivity analysis excluding four extreme p!zltau217 values (4–10 □pg/mL) did not alter the association between p-tau217 levels and dream recall (adjusted model: OR, 0.53; 95% CI, 0.35, 0.81; *p* = 0.003).

**Table 2.** Associations between p-tau217, *APOE* ε4 status, baseline cognition and dream recall.

| Characteristic | Unadjusted models |  |  | Adjusted models |  |  |
| --- | --- | --- | --- | --- | --- | --- |
|  | Coefficient | *p* value | OR (95% CI) | Coefficient | *p* value | OR (95% CI) |
| p-tau217, >0.247 pg/mL | -0.687 | 0.001 | 0.50 (0.34, 0.74) | -0.607 | 0.004 | 0.55 (0.36, 0.83) |
| *APOE* ε4 carrier | -0.456 | 0.008 | 0.63 (0.45, 0.89) | -0.482 | 0.007 | 0.62 (0.44, 0.87) |
| PACCm, z-score | 0.112 | 0.212 | 1.12 (0.94, 1.34) | 0.139 | 0.155 | 1.15 (0.95, 1.39) |

*Note:* Adjusted models included baseline covariates: age, sex, initial insomnia, maintenance insomnia, sleep duration, and awakenings per night. Free delayed recall score was also included when p-tau217 or *APOE* ε4 status was the predictor, and *APOE* ε4 status was included when p-tau217 or PACCm z-score was the predictor.
Abbreviations: *APOE*, apolipoprotein E; PACCm, modified Preclinical Alzheimer Cognitive Composite.

### 3.3 Association between dream recall and longitudinal cognitive trajectory

Linear mixed-effects model results show that cognitive scores declined over follow up and were lower for *APOE* ε4 carriers than for non-carriers (Table 3). There was a significant age by dream recall interaction, with cognitive decline twice as fast for non-dream recallers than for dream recallers (-0.030 vs -0.015 annually) (Figure 1).

**Table 3.** Association between dream recall and longitudinal trajectory of PACCm z-scores

| **Parameter** | **$\beta$ (95% CI)** | ***p* value** |
| --- | --- | --- |
| Intercept | 1.27 (0.79, 1.74) | <0.001 |
| Age | -0.02 (-0.02, -0.01) | <0.001 |
| Dream recall | 1.07 (0.27, 1.87) | 0.009 |
| Education | 0.07 (-0.03, 0.17) | 0.155 |
| Sex | -0.03 (-0.13, 0.07) | 0.559 |
| Age at baseline | -0.03 (-0.04, -0.01) | <0.001 |
| *APOE* ε4 carrier | -0.14 (-0.27, -0.02) | 0.028 |
| Age*Dream recall | -0.02 (-0.03, -0.01) | 0.005 |

*Note:* Age at baseline is centered at the median of 74 years.

**Figure 1.**
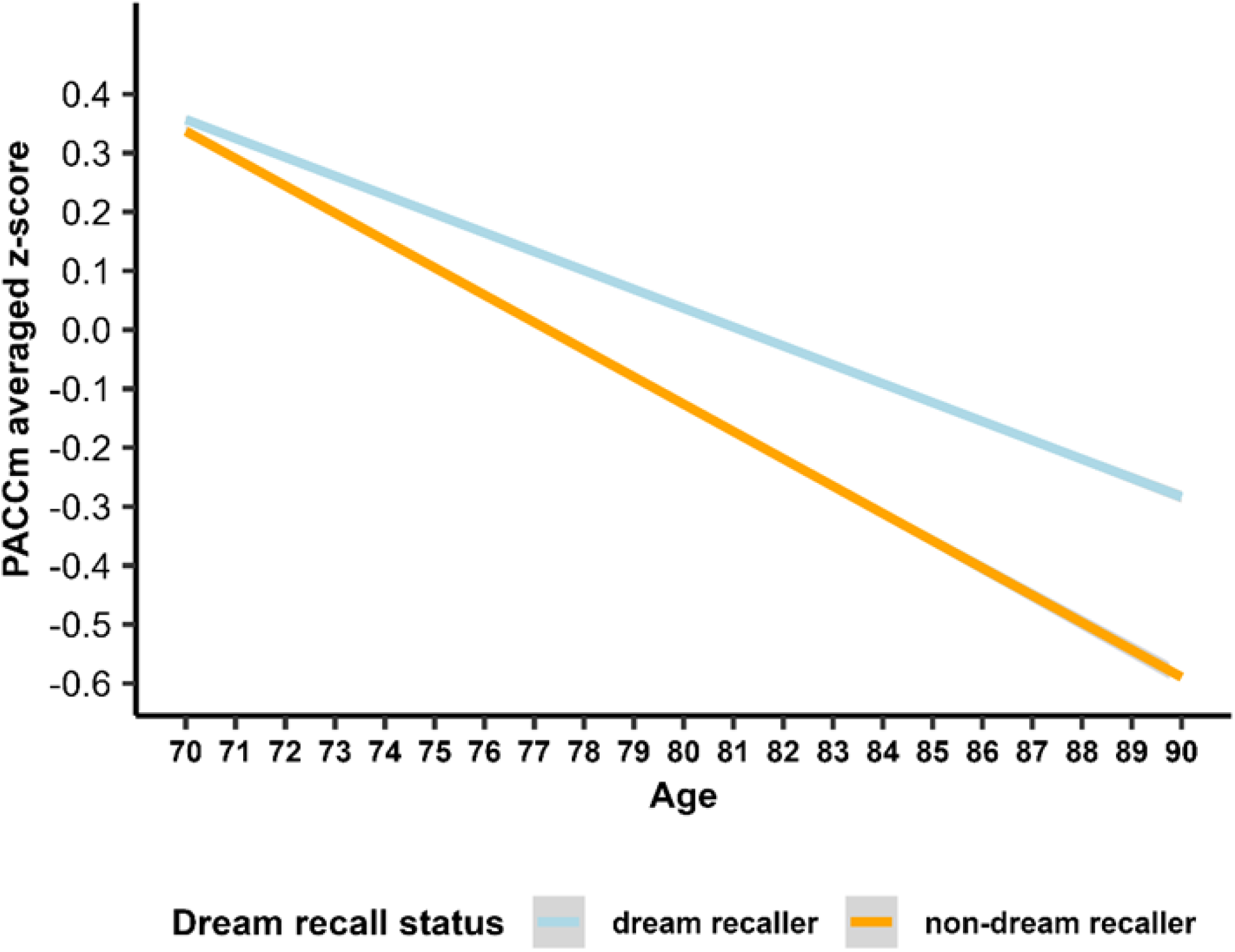
Cognitive trajectories of participants by dream recall status. The predicted regression lines represent the estimated values over time, from ages 70 to 90 y, for participants with primary or incomplete high school education, males, aged 74 y at baseline, and APOE _ε_4 non-carriers; differentiated by dream recall status: dream recallers (light blue) and non-recallers (orange).

### 3.4 Associations between dream recall and incident MCI and dementia

Univariate Cox regressions found increased risks of developing MCI and dementia for individuals who did not remember dreams, were an *APOE* e4 carrier, or older than 74 years (Table 4). After adjusting for age and sex, dream recall remained significantly associated with incident dementia, but only tended to be associated with incident MCI. In stratified analyses, the greater risk of dementia for non-recallers was maintained in *APOE* e4 carriers but not in non-carriers. Similar analyses for MCI found no significant effects in either *APOE* e4 carriers or non-carriers (Table 4).

**Table 4.** Associations between not recalling dreams and incident MCI and dementia.

| Characteristic | MCI |  |  | Dementia |  |  |
| --- | --- | --- | --- | --- | --- | --- |
|  | Coefficient | *p* value | HR (95% CI) | Coefficient | *p* value | HR (95% CI) |
| **Univariate models** |  |  |  |  |  |  |
| No dream recall | 0.32 | 0.025 | 1.38 (1.04, 1.83) | 0.51 | 0.027 | 1.67 (1.06, 2.60) |
| Sex, female | -0.02 | 0.899 | 0.98 (0.74, 1.30) | 0.34 | 0.176 | 1.41 (0.86, 2.31) |
| *APOE* ε4 carrier | 0.69 | <0.001 | 1.99 (1.46, 2.72) | 0.94 | <0.001 | 2.57 (1.6, 4.12) |
| Age at baseline | 0.08 | <0.001 | 1.08 (1.04, 1.12) | 0.07 | 0.021 | 1.08 (1.01, 1.10) |
| **No dream recall, adjusted models** |  |  |  |  |  |  |
| All participants | 0.27 | 0.057 | 1.32 (0.99, 1.70) | 0.49 | 0.038 | 1.62 (1.03, 2.60) |
| *APOE* ε4 non-carriers | 0.17 | 0.108 | 1.19 (0.85, 1.70) | 0.13 | 0.654 | 1.14 (0.64, 2.10) |
| *APOE* ε4 carriers | 0.45 | 0.312 | 1.57 (0.91, 2.70) | 1.10 | 0.007 | 3.04 (1.35, 6.90) |

*Note:* Age at baseline is centered at the median of 74 years. Adjusted models include baseline age and sex (male = 0, female = 1).
Abbreviations: *APOE*, apolipoprotein E; MCI, mild cognitive impairment.

## 4. DISCUSSION

On the basis of the DMN subserving dreaming, being impaired in *APOE* ε4 carriers, and showing pre-clinical AD-related neuropathology, we hypothesized that dream recall will be reduced in cognitively healthy older adults who are *APOE* ε4 carriers or positive for AD biomarkers, or who show future cognitive decline including dementia. The current study results support all these hypotheses. They are also consistent with the previous finding that lower dream recall frequency predicted future dementia,^4^ but go further by implicating early neurodegeneration and genetic risk in this effect.

While we found a clear association between dream recall and incident dementia, the association between dream recall and MCI was less emphatic, being significant in unadjusted analyses but only tending towards significance in adjusted analyses. This likely reflects MCI being a less clinically definitive endpoint than dementia, with a greater potential for misclassification^42^ and heterogeneous trajectories that can include long-term stability or reversion to normal cognition.^43^ Our additional finding that individuals not recalling dreams showed faster decline than recallers on a continuous measure of cognition avoids these issues and supports the association between not recalling dreams and future dementia. This association was found for the whole sample, but only for ε4 carriers in analyses stratified by *APOE* ε4 status. *APOE* ε4 carriers may be particularly susceptible to low dream recall and future dementia given their higher global Aβ burden and DMN tau levels^24,25^ and the potential for *APOE* ε4 to influence the DMN via non-amyloid pathways.^26^

The previously reported association between dream recall frequency and dementia was independent of subjectively assessed “memory for recent events”.^4^ The current study found that associations between dream recall and each of p-tau217 levels and *APOE* ε4 carriage were independent of objectively assessed memory test performance. Indeed, as found here and by others,^2,44^ memory performance seems to have no, or only a minimal, association with dream recall. Rather than reduced memory of dreams, poor dream recall may thus reflect an impaired dream production process consistent with the DMN being the neural substrate for dreaming,^8^ showing abnormal functionality in *APOE* ε4 carriers,^7,20,21,26^ and undergoing early Aβ and tau accumulation.^14^ The idea that DMN neuropathology can impair dreaming is supported by analogous effects in lesion and functional MRI studies.^8,9^

In addition to early abnormalities of DMN function, cognitively unimpaired *APOE* ε4 carriers have shown REM sleep duration.^45^ Lower REM sleep percentage and longer latency to REM from sleep onset have been associated with an increased risk of dementia, including AD,^46^ consistent with associations between prolonged REM latency and Aβ.^47^ This might suggest that *APOE* ε4 carriers and individuals with AD-neuropathology have reduced recall for dreams because of less REM sleep and therefore fewer dreams. Keeping in mind that the DMN was not yet discovered,^5^ less REM sleep was the tentative explanation for the previously observed association between lower dream recall frequency and higher risk of dementia.^4^ Conversely, amyloid positivity in cognitively unimpaired older adults has been associated with a reduction in REM sleep theta power.^48^ Another study associated higher REM sleep theta power with greater chances of dream recall.^49^ These results suggest that amyloid may contribute to impaired dream production during REM sleep. Further, less REM sleep does not account for impaired dreaming associated with focal cerebral lesions^50^ or surgically severed frontal lobe connections^51^ that leave REM sleep unaffected. Others have reported that the percentage of dreams recalled after laboratory awakenings from REM sleep was inversely associated with the degree of organicity among “chronic brain syndrome” patients.^52^ These findings are consistent with REM sleep and dreaming being separate processes regulated by brainstem and forebrain mechanisms, respectively.^50^ Reflecting this is the occurrence of dreaming in some non-REM sleep stages, though recall rates are lower than for REM sleep.^8,53^

While suggesting that impaired dream production may be implicated in our results, we cannot exclude effects associated with cognitive processes when awake. Evidence suggests that an awakening of at least two minutes is needed to encode a dream into long term memory.^54^ Attention could have a role in this process. Some trait attentional differences between high and low recallers during externally based attention tasks have been observed,^55,56^ but the implications for dream recall remain unclear. Encoding a dream upon awakening presumably involves attention to internally focused cognitions subserved by the DMN.^6,7^ It is thus not surprising that, compared to low recallers, high recallers showed greater functional connectivity in the DMN, and between the DMN and regions involved in memory, immediately after waking from an afternoon nap.^57^ Our results could therefore be at least partly explained by DMN dysfunction during waking rather than impaired dream production during sleep. Fisher-Hicks et al.^3^ gave a similar caveat regarding their recent findings of dream cessation after head injury. Of course, DMN dysfunction during both sleep and waking could be implicated in poor dream recall.

If DMN alterations associated with *APOE* ε4 carriage and/or AD related neuropathology do indeed contribute to poor dream recall, other DMN processes should show similar associations. Associations between proneness to mind wandering and remembering dreams was recently found, consistent with both being DMN processes.^2^ Decreased mind wandering in older adults should thus also be associated with p-tau217, *APOE* ε4 carriage, and cognitive decline. While a report of decreased mind-wandering in adults with mild dementia is ostensibly consistent with this, mind wandering was measured by asking participants about disengagement during a sustained attention task.^58^ Participants with dementia reported less disengagement, but also more interest in the task than healthy controls. A better measure to test our DMN hypothesis may be a propensity for daydreaming scale,^59^ for which scores have been associated with DMN activity during high-incidence periods of stimulus-independent thought.^60^

In addition to associations with p-tau217, *APOE* ε4 carriage and dementia, other factors were associated with dream recall in multivariate analyses. In line with intuition and previous studies, there were greater chances of dream recall associated with longer sleep^2,61,62^ and more overnight awakenings.^54,63^ Insomnia has been associated with both more^63^ and less^62^ dream recall. Our results suggest that the type of insomnia may be important, as initial and maintenance insomnia were associated with greater and lesser chances of dream recall, respectively. Evidence implicates hyperconnectivity and hyperactivation of the DMN in insomnia,^64,65^ suggesting a possible direct neurobiological link between insomnia and greater chances of dream recall. Conversely, smokers show reduced connectivity within the DMN,^66^ which could help explain our finding of less dream recall among smokers.

This study’s strengths include using data from a well-characterised cohort, allowing for the control of many extraneous factors potentially influencing dream recall, the long follow-up period, and the use of plasma p-tau217, which reportedly outperforms other plasma p-tau isoforms and has similar accuracy to CSF p-tau217.^27^ Limitation include the binary dream recall measure, as frequency data would allow “dose response” effects to be investigated, and not knowing how many dream recallers experienced nightmares, given that distressing dreams have been associated with an increased risk of dementia.^67^ The inclusion of nightmare experiencers in the dream recall group could mean our results underestimate the true effects. Lastly, we did not control for interest in or attitude towards dreams, which is known to be associated with greater recall, though the causal relationships are unclear.^2^

In conclusion, we found that dream recall status was associated with blood p-tau217 levels, *APOE* ε4 carriage, and future cognitive decline and dementia among older adults cognitively healthy at baseline. The association between dream recall and a strong genetic risk factor for AD (*APOE* ε4), as well as between dream recall and cognitive decline, dementia and an AD biomarker (p-tau217), can be considered a Mendelian randomization test corroborating the relationship between dream recall and AD. Our findings are consistent with the DMN’s role in dreaming, with impaired DMN functionality among *APOE* ε4 carriers, and with pre-clinical amyloid and tau deposition in the DMN. Further research is needed to directly determine whether altered DMN functionality is indeed implicated in our findings.

Nevertheless, if other factors affecting dream recall can be excluded, poor dream recall in later life may be an indicator of early neurodegeneration and increased risk of cognitive decline. With dreaming conceptualised as a form of internally generated thought,^8^ a self-reported loss of dream recall in cognitively normal adults could be considered analogous to subjective cognitive decline, which is thought to frequently be the first symptom of AD.^68^

## Data Availability

Anonymized data can be accessed upon request at.

## ACKNOWLEDGMENTS

We are indebted to all the volunteers who participated in the Vallecas Project.

## CONSENT STATEMENT

The Vallecas Project was approved by the Ethics Committee of the Carlos III Health Institute, and all participants provided informed consent according to the Declaration of Helsinki.

## REFERENCES

1. Van de Castle RL. Our dreaming mind. New York: Ballantine Books; 1994.

2. Elce V, Bergamo D, Bontempi G, et al. The individual determinants of morning dream recall. Commun Psychol. 2025;3(1):25.

3. Fisher-Hicks S, Lovett V, Wood RL, Blagrove M. The association of brain injury severity with dream cessation and nightmares. Neuropsychologia. 2025:109209.

4. Persson G, Skoog I. Subclinical dementia: relevance of cognitive symptoms and signs. J Geriatr Psychiatry Neurol. 1992;5(3):172–8.

5. Raichle ME, MacLeod AM, Snyder AZ, Powers WJ, Gusnard DA, Shulman GL. A default mode of brain function. Proc Natl Acad Sci U S A. 2001;98(2):676–82.

6. Menon V. 20 years of the default mode network: A review and synthesis. Neuron. 2023;111(16):2469–2487.

7. Mevel K, Chetelat G, Eustache F, Desgranges B. The default mode network in healthy aging and Alzheimer’s disease. Int J Alzheimers Dis. 2011;2011:535816.

8. Domhoff GW. The Neurocognitive Theory of Dreaming. Cambridge, Massachusetts: The MIT Press; 2022.

9. Eichenlaub JB, Nicolas A, Daltrozzo J, Redoute J, Costes N, Ruby P. Resting brain activity varies with dream recall frequency between subjects. Neuropsychopharmacology. 2014;39(7):1594–602.

10. Vallat R, Eichenlaub JB, Nicolas A, Ruby P. Dream Recall Frequency Is Associated With Medial Prefrontal Cortex White-Matter Density. Front Psychol. 2018;9:1856.

11. Vallat R, Turker B, Nicolas A, Ruby P. High Dream Recall Frequency is Associated with Increased Creativity and Default Mode Network Connectivity. Nat Sci Sleep. 2022;14:265–275.

12. Zhang Q, Li F, Wei M, et al. Prediction of Cognitive Progression Due to Alzheimer’s Disease in Normal Participants Based on Individual Default Mode Network Metabolic Connectivity Strength. Biol Psychiatry Cogn Neurosci Neuroimaging. 2024;9(7):660–667.

13. Ereira S, Waters S, Razi A, Marshall CR. Early detection of dementia with default-mode network effective connectivity. Nature Mental Health. 2024;2:787–800.

14. Palmqvist S, Scholl M, Strandberg O, et al. Earliest accumulation of beta-amyloid occurs within the default-mode network and concurrently affects brain connectivity. Nat Commun. 2017;8(1):1214.

15. Ingala S, Tomassen J, Collij LE, et al. Amyloid-driven disruption of default mode network connectivity in cognitively healthy individuals. Brain Commun. 2021;3(4):fcab201.

16. Schultz AP, Chhatwal JP, Hedden T, et al. Phases of Hyperconnectivity and Hypoconnectivity in the Default Mode and Salience Networks Track with Amyloid and Tau in Clinically Normal Individuals. J Neurosci. 2017;37(16):4323–4331.

17. Sepulcre J, Sabuncu MR, Li Q, El Fakhri G, Sperling R, Johnson KA. Tau and amyloid beta proteins distinctively associate to functional network changes in the aging brain. Alzheimers Dement. 2017;13(11):1261–1269.

18. Putcha D, Eckbo R, Katsumi Y, Dickerson BC, Touroutoglou A, Collins JA. Tau and the fractionated default mode network in atypical Alzheimer’s disease. Brain Commun. 2022;4(2):fcac055.

19. Katsumi Y, Howe IA, Eckbo R, et al. Default mode network tau predicts future clinical decline in atypical early Alzheimer’s disease. Brain. 2024;

20. Kuang L, Jia J, Zhao D, et al. Default Mode Network Analysis of APOE Genotype in Cognitively Unimpaired Subjects Based on Persistent Homology. Front Aging Neurosci. 2020;12:188.

21. Westlye ET, Lundervold A, Rootwelt H, Lundervold AJ, Westlye LT. Increased hippocampal default mode synchronization during rest in middle-aged and elderly APOE epsilon4 carriers: relationships with memory performance. J Neurosci. 2011;31(21):7775–83.

22. Corder EH, Saunders AM, Strittmatter WJ, et al. Gene dose of apolipoprotein E type allele and the risk of Alzheimer’s disease in late onset families. Science. 1993;261(5123):921–3.

23. Liu CC, Liu CC, Kanekiyo T, Xu H, Bu G. Apolipoprotein E and Alzheimer disease: risk, mechanisms and therapy. Nat Rev Neurol. 2013;9(2):106–18.

24. Therriault J, Benedet AL, Pascoal TA, et al. Association of Apolipoprotein E epsilon4 With Medial Temporal Tau Independent of Amyloid-beta. JAMA Neurol. 2020;77(4):470–479.

25. Young CB, Johns E, Kennedy G, et al. APOE effects on regional tau in preclinical Alzheimer’s disease. Mol Neurodegener. 2023;18(1):1.

26. Sheline YI, Morris JC, Snyder AZ, et al. APOE4 allele disrupts resting state fMRI connectivity in the absence of amyloid plaques or decreased CSF Abeta42. J Neurosci. 2010;30(50):17035–40.

27. Lai R, Li B, Bishnoi R. P-tau217 as a Reliable Blood-Based Marker of Alzheimer’s Disease. Biomedicines. 2024;12(8)

28. Putois B, Leslie W, Asfeld C, Sierro C, Higgins S, Ruby P. Methodological Recommendations to Control for Factors Influencing Dream and Nightmare Recall in Clinical and Experimental Studies of Dreaming. Front Neurol. 2020;11:724.

29. Olazaran J, Valenti M, Frades B, et al. The Vallecas Project: A Cohort to Identify Early Markers and Mechanisms of Alzheimer’s Disease. Front Aging Neurosci. 2015;7:181.

30. Calero O, Hortiguela R, Bullido MJ, Calero M. Apolipoprotein E genotyping method by real time PCR, a fast and cost-effective alternative to the TaqMan and FRET assays. J Neurosci Methods. 2009;183(2):238–40.

31. Tanne JH. FDA approves blood test to diagnose Alzheimer’s. BMJ. 2025;389:r1082.

32. Arranz J, Zhu N, Rubio-Guerra S, et al. Diagnostic performance of plasma pTau(217), pTau(181), Abeta(1-42) and Abeta(1-40) in the LUMIPULSE automated platform for the detection of Alzheimer disease. Alzheimers Res Ther. 2024;16(1):139.

33. Langbaum JB, Ellison NN, Caputo A, et al. The Alzheimer’s Prevention Initiative Composite Cognitive Test: a practical measure for tracking cognitive decline in preclinical Alzheimer’s disease. Alzheimers Res Ther. 2020;12(1):66.

34. Buschke H. Cued recall in amnesia. J Clin Neuropsychol. 1984;6(4):433–40.

35. Wechsler D. Wechsler Adult Intelligence Scale-III. San Antonio: The psychological Corporation; 1997.

36. Folstein MF, Folstein SE, McHugh PR. “Mini-mental state”. A practical method for grading the cognitive state of patients for the clinician. J Psychiatr Res. 1975;12(3):189–98.

37. Winblad B, Palmer K, Kivipelto M, et al. Mild cognitive impairment--beyond controversies, towards a consensus: report of the International Working Group on Mild Cognitive Impairment. J Intern Med. 2004;256(3):240–6.

38. Pfeffer RI, Kurosaki TT, Harrah CH, Jr., Chance JM, Filos S. Measurement of functional activities in older adults in the community. J Gerontol. 1982;37(3):323–9.

39. American Psychiatric Association. Diagnostic and statistical manual of mental disorders (4th ed., text rev.) Washington, DC: American Psychiatric Association,; 2000.

40. Sheikh JI, Yesavage JA. Recent evidence and development of a shorter version. Clinical Gerontologist: The Journal of Aging and Mental Health. 1986;5(1-2):165–173.

41. Spielberger CD, Gorsuch RL, Lushene RE. Manual for the State-Trait Anxiety Inventory (STAI). Palo Alto, CA: Consulting Psychologists Press; 1970.

42. Brooks BL, Iverson GL, Holdnack JA, Feldman HH. Potential for misclassification of mild cognitive impairment: a study of memory scores on the Wechsler Memory Scale-III in healthy older adults. J Int Neuropsychol Soc. 2008;14(3):463–78.

43. Salemme S, Lombardo FL, Lacorte E, et al. The prognosis of mild cognitive impairment: A systematic review and meta-analysis. Alzheimers Dement (Amst). 2025;17(1):e70074.

44. Waterman D. Aging and memory for dreams. Percept Mot Skills. 1991;73(2):355–65.

45. Andre C, Martineau-Dussault ME, Baril AA, et al. Reduced rapid eye movement sleep in late middle-aged and older apolipoprotein E ε4 allele carriers. Sleep. 2024;47(7)

46. Pase MP, Himali JJ, Grima NA, et al. Sleep architecture and the risk of incident dementia in the community. Neurology. 2017;89(12):1244–1250.

47. Jin J, Chen J, Cavailles C, et al. Association of rapid eye movement sleep latency with multimodal biomarkers of Alzheimer’s disease. Alzheimers Dement. 2025;21(2):e14495.

48. Andre C, Champetier P, Rehel S, et al. Rapid Eye Movement Sleep, Neurodegeneration, and Amyloid Deposition in Aging. Ann Neurol. 2023;93(5):979–990.

49. Marzano C, Ferrara M, Mauro F, et al. Recalling and forgetting dreams: theta and alpha oscillations during sleep predict subsequent dream recall. J Neurosci. 2011;31(18):6674–83.

50. Solms M. Dreaming and REM sleep are controlled by different brain mechanisms. Behav Brain Sci. 2000;23(6):843-50; discussion 904-1121.

51. Jus A, Jus K, Villeneuve A, et al. Studies on dream recall in chronic schizophrenic patients after prefrontal lobotomy. Biol Psychiatry. 1973;6(3):275–93.

52. Kramer M, Roth T. Dreams and dementia: a laboratory exploration of dream recall and dream content in chronic brain syndrome patients. Int J Aging Hum Dev. 1975;6(2):179–82.

53. Nielsen TA. A review of mentation in REM and NREM sleep: “covert” REM sleep as a possible reconciliation of two opposing models. Behav Brain Sci. 2000;23(6):851-66; discussion 904-1121.

54. Vallat R, Lajnef T, Eichenlaub JB, et al. Increased Evoked Potentials to Arousing Auditory Stimuli during Sleep: Implication for the Understanding of Dream Recall. Front Hum Neurosci. 2017;11:132.

55. Blain S, de la Chapelle A, Caclin A, Bidet-Caulet A, Ruby P. Dream recall frequency is associated with attention rather than with working memory abilities. J Sleep Res. 2022;31(5):e13557.

56. Ruby P, Masson R, Chatard B, et al. High dream recall frequency is associated with an increase of both bottom-up and top-down attentional processes. Cereb Cortex. 2022;32(17):3752–3762.

57. Vallat R, Nicolas A, Ruby P. Brain functional connectivity upon awakening from sleep predicts interindividual differences in dream recall frequency. Sleep. 2020;43(12)

58. Gyurkovics M, Balota DA, Jackson JD. Mind-wandering in healthy aging and early stage Alzheimer’s disease. Neuropsychology. 2018;32(1):89–101.

59. Giambra LM. The influence of aging on spontaneous shifts of attention from external stimuli to the contents of consciousness. Exp Gerontol. 1993;28(4-5):485-92.

60. Mason MF, Norton MI, Van Horn JD, Wegner DM, Grafton ST, Macrae CN. Wandering minds: the default network and stimulus-independent thought. Science. 2007;315(5810):393–5.

61. Schredl M, Reinhard I. Dream recall, dream length, and sleep duration: state or trait factor. Percept Mot Skills. 2008;106(2):633–6.

62. Tschunichin D, Schredl M. Dream recall, white dreaming, and sleep duration: A diary study in patients with sleep disorders. Somnologie. 2024;

63. Schredl M, Schafer G, Weber B, Heuser I. Dreaming and insomnia: dream recall and dream content of patients with insomnia. J Sleep Res. 1998;7(3):191–8.

64. Schiel JE, Holub F, Petri R, et al. Affect and Arousal in Insomnia: Through a Lens of Neuroimaging Studies. Curr Psychiatry Rep. 2020;22(9):44.

65. Zhang L, Guo Z, Han Y, et al. Altered cerebral functional activity and its associated genetic profiles underlying chronic insomnia disorder before and after treatment. World J Biol Psychiatry. 2025;26(5):211–223.

66. Weiland BJ, Sabbineni A, Calhoun VD, Welsh RC, Hutchison KE. Reduced executive and default network functional connectivity in cigarette smokers. Hum Brain Mapp. 2015;36(3):872–82.

67. Otaiku AI. Distressing dreams, cognitive decline, and risk of dementia: A prospective study of three population-based cohorts. EClinicalMedicine. 2022;52:101640.

68. Jessen F, Amariglio RE, van Boxtel M, et al. A conceptual framework for research on subjective cognitive decline in preclinical Alzheimer’s disease. Alzheimers Dement. 2014;10(6):844–52.

