## Supplementary material for "Poor dream recall associates with Alzheimer’s disease biomarkers and dementia risk": Table S1

**Table S1.** Median (IQR) for continuous variables at baseline

| **Characteristic** | **Total** |  | **Dream recall status** | |
| --- | --- | --- | --- | --- |
|  |  |  | **Yes** | **No** |
| Age, years | 74 (5) |  | 74 (6) | 74 (6) |
| p-tau217, pg/mL | 0.107 (0.097) |  | 0.102 (0.085) | 0.121 (0.130) |
| Total delayed recall | 15 (2) |  | 15 (2) | 15 (2) |
| Free delayed recall | 10 (3) |  | 10 (3) | 9 (3) |
| PACCm, z-scores | 0.03 (1.0) |  | 0.05 (1.1) | -0.02 (1.0) |
| Awakenings per night | 1 (1) |  | 1 (1.3) | 1 (2) |
| Parkinsonian symptoms | 0 (0) |  | 0 (0) | 0 (0) |
| GDS score | 1 (2) |  | 1 (2) | 1 (2) |
| State anxiety score | 13 (10) |  | 13 (10) | 13 (10) |
| Trait anxiety score | 15 (14) |  | 16 (13.5) | 14 (13) |
